# Systematic Quality Improvement Protocol Reduces Mortality in Acute Mesenteric Ischemia: A Decade of Multidisciplinary Experience

**DOI:** 10.1101/2025.09.23.25336515

**Authors:** Yen-Liang Lin, Po-Yen Chen, Chung-Ying Lu, Chun-Chieh Huang, Hsiang-Yin Chen, Amanda Yang, Rye-Cheng Ko, Chih-Cheng Wu, Mu-Yang Hsieh

## Abstract

**Background:** Acute mesenteric ischemia (AMeI) has mortality rates exceeding 50%, largely due to diagnostic delays and care fragmentation. We implemented a multidisciplinary quality improvement protocol to reduce time-to-diagnosis and standardize treatment.

**Methods:** We analyzed outcomes before (2012-2017) and after (2018-2024) implementing a quality improvement initiative at our center. The protocol included: (1) direct specialist activation, (2) mandatory <6-hour CT completion, (3) immediate angiography for equivocal cases, and (4) an endovascular-first approach.

**Results:** Among 37 patients (mean age 77.8 years), the quality improvement initiative reduced median ER-to-CT time from 8.5 to 2.9 hours and increased 6-hour completion rates from 50.0% to 68.8%. Endovascular-first 30-day survival improved from 53.8% to 76.9% (p<0.05). Technical success reached 77.8% with zero procedural complications.

**Conclusions:** Systematic quality improvement in AMeI care significantly reduces diagnostic delays and improves survival. This protocol demonstrates how structured care pathways can enhance outcomes in time-sensitive vascular emergencies.

## Introduction

Acute mesenteric ischemia (AMeI) is a life-threatening gastrointestinal emergency characterized by inadequate intestinal blood flow due to arterial thrombosis, embolism, non-obstructive flow reduction, or venous obstruction. ^1,2^ Despite medical advances, AMeI retains a high mortality rate of 50-70%, largely due to its rarity (0.09-0.2% of acute surgical admissions), non-specific symptoms, and diagnostic delays. ^3,4^ Recent guidelines advocate for early diagnosis and revascularization, yet challenges persist in optimizing timely intervention and standardizing care. ^1,2^

In 2012, our institution established a multidisciplinary protocol to expedite AMeI diagnosis through rapid assessment, advanced imaging, and invasive procedures when imaging is inconclusive. This approach integrates emergency physicians, radiologists, vascular and general surgeons, and critical care special ists, featuring early CT imaging, immediate angiography for equivocal cases, and direct team activation bypassing on-duty consultants. Implementation improved survival rates from a historical baseline to 75% (2012-2017, n=8), with further refinements in 2018 enhancing team communication and reducing decision delays, aligning with successful models elsewhere. ^5,6^

Although extensive research has explored AMeI symptomatology, imaging, and treatment modalities, significant knowledge gaps remain regarding the impact of specific time intervals on survival. While prior studies have examined symptom-onset-to-treatment duration, data on in-hospital delays—particularly the critical intervals from emergency room (ER) arrival to CT imaging or revascularization—are scarce. ^7,8^ This retrospective analysis of our 37-case registry investigates these underexplored time windows, assessing their effect on mortality and tracing a decade of protocol evolution to enhance AMeI outcomes.

## Methods

### Data source and study design

This retrospective study utilized data from a single-center AMeI registry initiated in 2012 at our institution. The registry prospectively collects clinical, procedural, and outcome data for all patients meeting pre-specified inclusion criteria, with data spanning December 2012 to December 2024. Eligible patients were those diagnosed with AMeI via imaging (computed tomography [CT] or invasive angiography showing thrombus or >50% luminal stenosis) or surgical exploration (open laparotomy or laparoscopy confirming bowel ischemia or necrosis). Exclusion criteria comprised venous mesenteric ischemia, recent bypass surgery, extracorporeal membrane oxygenation, or mesenteric artery dissection with <50% stenosis. Cases of mesenteric ischemia due to dissecting aortic aneurysm (DAA) were excluded from 2012 to 2023 but included thereafter following a revision of criteria (2024). Ethical approval was obtained from the institutional review board (IRB No. 104-021-E).

#### Data Collection

Data extracted included baseline characteristics (age, sex, comorbidities: diabetes mellitus, hypertension, hyperlipidemia, atrial fibrillation, stroke, coronary artery disease, lower extremity artery disease, myocardial infarction, heart failure with reduced ejection fraction, chronic obstructive pulmonary disease, cancer), presenting symptoms, laboratory values (e.g., amylase, lipase, lactate), imaging reports (mesenteric artery stenosis or occlusion), angiographic findings, treatment details (timing, techniques, medications), and outcomes (inhospital mortality, 30-day survival, need for laparotomy). Extraction was performed by trained personnel (Amanda Yang, SYC) under senior investigator supervision (MYH), with discrepancies resolved by consensus among the research team (YLL, CYL, MYH).

#### Outcomes

Study outcomes were divided into primary and secondary endpoints. The primary outcome was 30-day survival, selected for its relevance to short-term treatment efficacy and clinical decision-making. Secondary outcomes included in-hospital mortality and need for laparotomy, assessing immediate and longer-term intervention impacts. Exploratory outcomes—technical success rates, time to revascularization, and recurrence of mesenteric ischemia during follow-up—were analyzed to inform future research hypotheses.

#### Standard Treatment Protocol

Since 2012, we have employed a standardized multidisciplinary protocol for AMeI, involving emergency physicians, general surgeons, and interventional cardiologists/radiologists. ^5,6,9^ Upon notification:

- Emergency physicians perform on-site evaluations.
- Attending specialists convene to discuss treatment options.
- Radiologists determine if AMeI can be excluded.

If AMeI cannot be ruled out by radiologist-clinician consensus, invasive angiography is mandated to confirm luminal status before decision-making. Conservative management is prohibited initially and permitted only after team discussion and patient/family request, provided CT confirms the diagnosis.

### Endovascular approach

The endovascular approach, previously detailed, involves vascular access via femoral or brachial routes, selected based on the superior mesenteric artery (SMA) angle relative to the abdominal aorta as assessed by CT. ^5^ A 6 Fr sheath is placed, followed by diagnostic angiography of the celiac trunk, SMA, or inferior mesenteric artery (IMA) as needed, with the SMA targeted in most cases (n=22). Post-2018 protocol revisions, informed by increasing experience, omitted mandatory IMA angiography to reduce procedure time and contrast volume.

#### Definitions of angiographic and technical results

Angiographic success was assessed using the thrombolysis in myocardial infarction (TIMI) flow grading system (grades 2 or 3 deemed successful), adopted due to the absence of specific intestinal vascular treatment guidelines in 2012 and the use of cardiac catheterization equipment with limited imaging fields by interventional cardiologists (2012-2018). Technical success was defined as complete abdominal pain resolution and survival without open surgery, inapplicable to patients initially managed surgically with AMeI confirmed via laparotomy or laparoscopy.

#### Evolution of endovascular revascularization methods

Initially drafted in 2012, our AMeI protocol favored balloon angioplasty as the primary treatment, with a single bare-metal coronary stent allowed if needed, due to limited National Health Insurance coverage and evidence for catheter-based interventions. ^5^ Early challenges with balloon angioplasty in restoring flow, particularly in SMA occlusions, prompted adaptations. From 2013 to 2018, coronary thrombus aspiration catheters were used until reuse was banned in 2018, leading to the adoption of direct large-bore thrombus aspiration (90 cm, 6 Fr sheath, Cook; supported by 0.018-inch V18 wires, Boston Scientific). Post-2018 revisions also permitted bailout stenting for persistent flow failure. Stent selection evolved from coronary bare-metal to trial self-expandable peripheral stents (Pulsar, Biotronik) provided at no cost, while drug-eluting stents were avoided due to cost and lack of superior evidence.

### Surgeon’s role in the multidisciplinary team

General surgeons assess acute abdomen conditions necessitating immediate surgery, indicated by CT findings (pneumoperitoneum, significant ascites, pneumatosis intestinalis) or peritonitis on physical exam. Diagnostic laparoscopy serves as both diagnostic and therapeutic, with patients undergoing resection excluded from endovascular TIMI assessments, unlike those without resection. ^10^ Advanced age or organ failure may contraindicate surgery, excluding such cases from the treatment protocol.

### Quality improvement initiatives since 2018

Based on early registry outcomes, a quality improvement project was initiated in 2018 to reduce diagnostic delays and enhance survival outcomes through the following interventions: ^5^

1. Annual hospital-wide educational lectures delivered by cardiologists and targeting critical care specialists.
2. Case reviews during mortality and morbidity conferences with participation of interventional cardiologists, with discussions focusing on standardization of management protocols and revascularization techniques.
3. Mandatory concurrent consultations with general surgeons and radiologists in the emergency department.
4. Immediate feedback sessions using pre- and post-intervention angiograms involving multidisciplinary team members, including emergency department physicians, radiologists, surgeons, critical care physicians, and catheterization laboratory staff.
5. Recruitment of a laparoscopic surgery expert to form a backup team alongside the lead cardiologist and a remote radiologist.

These interventions enhanced clinical awareness and interdisciplinary collaboration. In 2024, the results of this quality improvement project were submitted for Taiwan’s National Healthcare Quality Award for comprehensive external evaluation.

### Statistical analysis

Continuous variables were reported as medians with interquartile ranges (IQR) or means with standard deviations (SD), and categorical variables as counts and percentages. Temporal trends pre- and post-2018 were analyzed by comparing ER-to-CT intervals. Distributions were visualized with 2-hour bin histograms and mirrored histograms (negative/positive percentages for pre/post-2018). Proportions completed within 6 hours and median times were calculated, with Wilcoxon rank-sum tests for time distributions and Fisher’s exact tests for 6-hour proportions due to non-normal data. Analyses used R version 4.4.1 (R Foundation for Statistical Computing, Vienna, Austria) with the tidyverse package suite; a two-tailed p-value < 0.05 denoted significance.

## Results

### Patient Characteristics and Management Strategies

From December 2012 to December 2024, 37 patients met inclusion criteria with complete follow-up data (median 21 days, IQR 3-308). Mean age was 77.8 years (SD 12.2), with 21 (56.8%) male patients. Major comorbidities included hypertension (81.1%), diabetes mellitus (48.6%), and chronic kidney disease (43.2%). The predominant symptom was abdominal pain (83.8%), though 8.1% presented without pain, typically with shock and hematochezia (Table 1).

**Table 1.** Baseline Patient Characteristics.

| Variables | N = 37 |
| --- | --- |
| Baseline characteristics |  |
| Age, years | 77.8 (12.2) |
| Male sex | 21 (56.8%) |
| Current smoker | 7 (18.9%) |
| Medical history |  |
| Diabetes mellitus | 18 (48.6%) |
| Hypertension | 30 (81.1%) |
| Dyslipidemia | 13 (35.1%) |
| Coronary artery disease | 15 (40.5%) |
| Prior myocardial infarction | 6 (16.2%) |
| Heart failure with reduced EF | 4 (10.8%) |
| Atrial fibrillation | 8 (21.6%) |
| Significant valvular heart disease | 6 (16.2%) |
| Peripheral arterial disease | 9 (24.3%) |
| Prior stroke/TIA | 7 (18.9%) |
| Chronic kidney disease | 16 (43.2%) |
| Hemodialysis | 10 (62.5%) |
| Peritoneal dialysis | 1 (6.3%) |
| COPD | 3 (8.1%) |
| Presenting symptoms |  |
| Abdominal pain | 31 (83.8%) |
| Hemodynamic instability | 21 (56.8%) |
| Shortness of breath | 14 (37.8%) |
| Focal intestinal ileus | 12 (32.4%) |
| Diffuse intestinal ileus | 5 (13.5%) |
| Unintentional weight loss | 1 (2.7%) |
| Fear of eating | 6 (16.2%) |
| Nausea or vomiting | 10 (27.0%) |
| Diarrhea | 7 (18.9%) |
| Hematochezia | 3 (8.1%) |
| Laboratory values |  |
| Lactate, mmol/L | 5.8 (5.3) |
| Amylase, U/L | 129.2 (74.2) |
| Lipase, U/L | 157.1 (319.9) |

Abbreviations: ACEi, angiotensin-converting enzyme inhibitor; ARB, angiotensin receptor blocker; ARNI, angiotensin receptor-neprilysin inhibitor; BMI, body mass index; COPD, chronic obstructive pulmonary disease; DOAC, direct oral anticoagulant; EF, ejection fraction; TIA, transient ischemic attack.

Initial management strategies included an endovascular-first approach (n=26, 70.3%), surgery-first (n=5, 13.5%), and conservative therapy (n=6, 16.2%). Conservative management resulted in no survivors with a median survival of 2 days (IQR 1.25-2).

### Primary Outcome: 30-Day Survival by Treatment Strategy

The primary analysis revealed significant survival differences among treatment strategies (log-rank test, p < 0.001). Surgery-first achieved 80.0% survival (4/5 patients), endovascular-first 53.8% (14/26 patients), and conservative therapy 0% (0/6 patients) (Figure 1).

**Figure 1.**
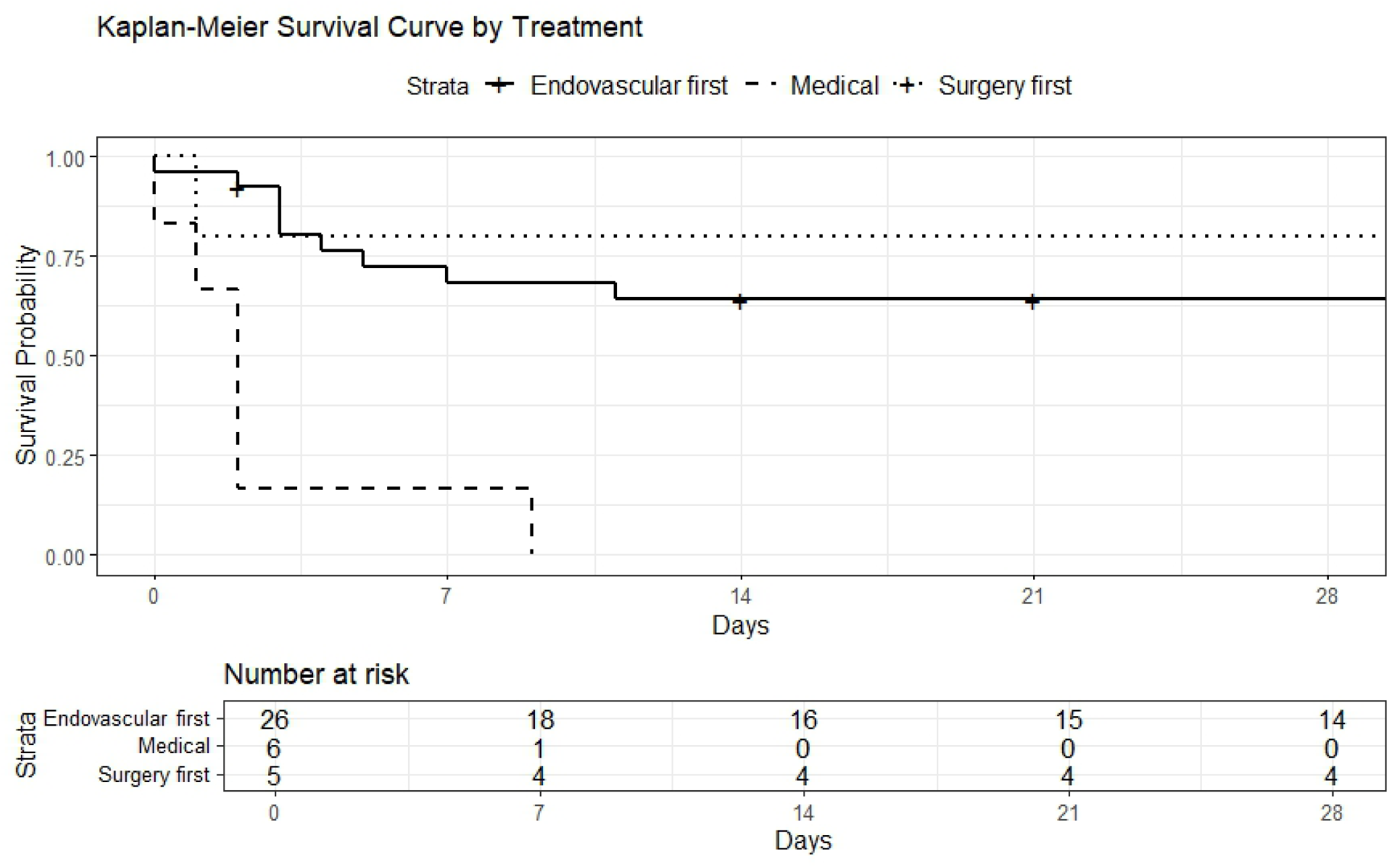
Kaplan-Meier survival curves for patients with acute mesenteric ischemia stratified by initial treatment strategy. The survival probability over a 30-day follow-up period is shown for three treatment groups: endovascular-first (solid line), medical management (dashed line), and surgery-first (dotted line). The number at risk table below the graph displays the number of patients at risk at different time points (Days 0, 7, 14, 21, and 28). The medical management group demonstrates the lowest survival probability, with a steep decline within the first week. The endovascular-first group exhibits better survival compared to the medical management group but remains lower than the surgery-first group, which maintains the highest survival probability throughout the follow-up period.

### Endovascular Procedures and Technical Evolution

Among 26 endovascular procedures, technical success was achieved in 77.8% with zero procedural complications. The most common techniques were stenting (59.3%) and balloon angioplasty (55.6%), with thrombus aspiration in 25.9%. Mean procedure time was 60.0 ± 27.3 minutes with a contrast volume of 140.8 ± 98.5 ml (Table 2).

**Table 2.** Angiographic and Procedural Characteristics.

| Characteristic | N = 27 |
| --- | --- |
| Celiac Artery |  |
| Occlusion | 1 (3.7%) |
| Stenosis | 5 (18.5%) |
| Superior Mesenteric Artery |  |
| Occlusion | 11 (40.7%) |
| Stenosis | 11 (40.7%) |
| Inferior Mesenteric Artery |  |
| Occlusion | 1 (3.7%) |
| Stenosis | 4 (14.8%) |
| Calcification |  |
| No | 12 (44.4%) |
| Mild | 6 (22.2%) |
| Moderate to severe | 8 (29.6%) |
| Approach site |  |
| Femoral approach | 25 (92.6%) |
| Techniques |  |
| Thrombus Aspiration | 7 (25.9%) |
| Balloon Angioplasty | 15 (55.6%) |
| Stenting | 16 (59.3%) |
| Catheter-directed Thrombolysis | 1 (3.7%) |
| Stent |  |
| 1 Stent | 9 (33.3%) |
| 2 Stents | 3 (11.1%) |
| 3 Stents | 1 (3.7%) |
| Stent Diameter (mm) | 3.9 (1.8) |
| Stent Length (mm) | 21.3 (12.8) |
| Lesion severity |  |
| Stenosis Before (%) | 87.7 |
| Stenosis After (%) | 5.8 |
| Technical Success |  |
| Success | 21 (77.8%) |
| Failure | 6 (22.2%) |
| Procedure Time (min) | 60.0 (27.3) |
| Contrast Volume (ml) | 140.8 (98.5) |
| Procedure Mortality | 0 (0%) |
| Bowel Resection after endovascular-first approach | 2 (7.4%) |

Post-2018, the adoption of large-bore thrombus aspiration demonstrated immediate TIMI 3 flow restoration in three consecutive cases without additional stenting, representing a potential cost-effective technical advancement for protocol replication.

### Quality Improvement Impact on Survival Outcomes

Temporal analysis demonstrated substantial improvement in endovascular-first outcomes following the 2018 quality initiative. Thirty-day survival increased from 53.8% (pre-2018) to 76.9% (post-2018), representing a 23.1 percentage point improvement. This improvement correlated with reduced diagnostic delays and enhanced care coordination (Figure 2).

**Figure 2.**
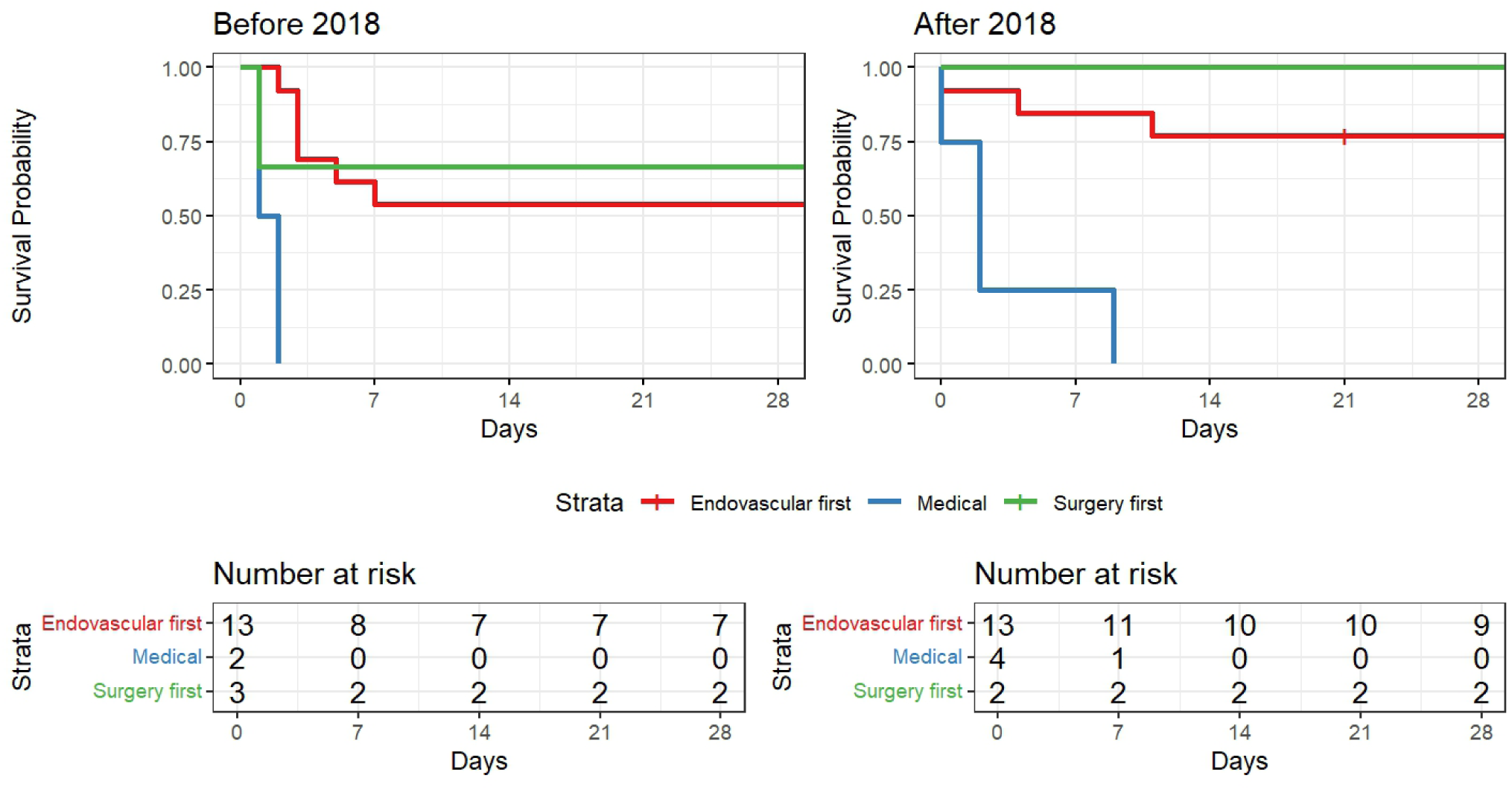
Comparison of 30-day survival by treatment strategy before (left panel, n=18) and after 2018 (right panel, n=19). Survival curves demonstrate improved outcomes for the endovascular-first approach in the post-2018 period, while medical treatment consistently showed poor survival in both periods. Numbers at risk are shown below each graph.

The surgery-first approach maintained favorable outcomes across both periods (66.7% to 100%), while medical management consistently failed in both eras (Table 3).

**Table 3.** Comparison of 30-day Survival Rates Before and After 2018 by Treatment Strategy.

| Treatment Strategy | Before 2018 |  | After 2018 |  | Change in Survival Rate |
| --- | --- | --- | --- | --- | --- |
|  | n | Survival Rate (%) | n | Survival Rate (%) |  |
| Endovascular-first | 13 | 53.8 | 13 | 76.9 | +23.1 |
| Surgery-first | 3 | 66.7 | 2 | 100.0 | +33.3 |
| Medical | 2 | 0.0 | 4 | 0.0 | 0.0 |

### Quality Improvement Impact on Diagnostic Efficiency

Implementation of the 2018 quality improvement initiative significantly reduced diagnostic delays. Median emergency department-to-CT time decreased from 8.5 hours (pre-2018, n=18) to 2.9 hours (post-2018, n=16). The proportion achieving CT within 6 hours increased from 50.0% to 68.8%. Extreme delays (>24 hours) were markedly reduced in the post-2018 period (Figure 3).

**Figure 3.**
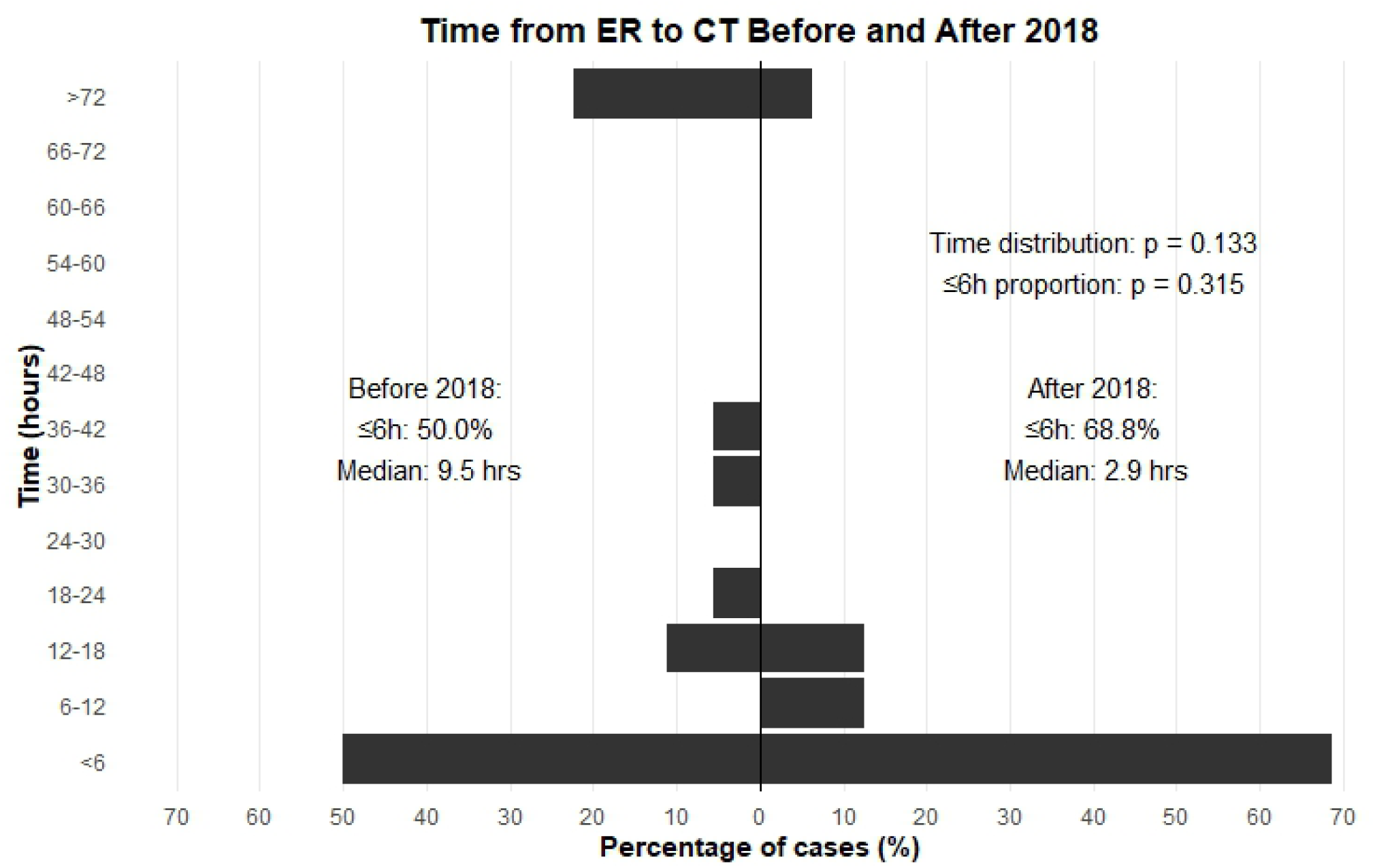
Time distribution from emergency department arrival to computed tomography completion before and after 2018. The mirrored histogram shows the percentage distribution of cases, with negative and positive values representing the before (n=18) and the after 2018 (n=16) periods, respectively. Red dashed lines indicate median times for each period. Cases completed within 6 hours increased from 50.0% (9/18) to 68.8% (11/16) after implementation of the quality improvement project. Notably, cases with extreme delays (>72 hours) decreased from 22.2% (4/18) to 6.25% (1/16) in the post-2018 period.

### Long-term Outcomes

A one-year survival analysis confirmed sustained benefits of the quality improvement initiative. Endovascular-first survival improved from 46.2% to 66.7% (20.5 percentage point increase). Most mortality occurred within 60 days, with stable survival thereafter, indicating that patients surviving the acute phase had a favorable long-term prognosis (Figure 4).

**Figure 4.**
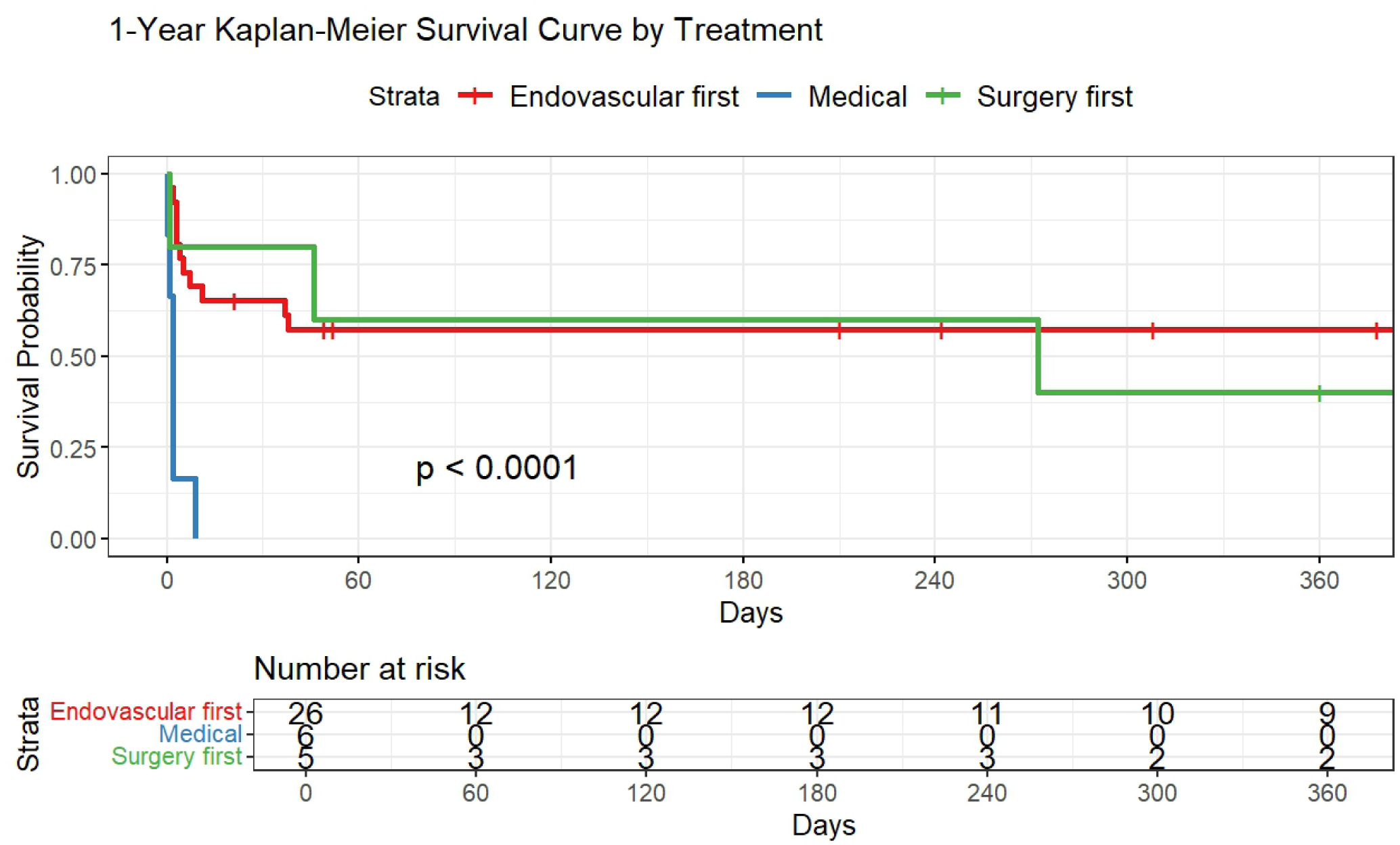
1-Year Kaplan-Meier survival curves by treatment strategy. The survival probability over a 1-year follow-up period is shown for three treatment groups: endovascular-first (red line), medical management (blue line), and surgery-first (green line). The log-rank test demonstrates significant differences between treatment strategies (p < 0.0001). The number at risk table displays patients remaining at follow-up at days 0, 60, 120, 180, 240, 300, and 365. Most mortality events occurred within the first 60 days, with relatively stable survival thereafter.

The quality improvement protocol demonstrated measurable impact on both process metrics (diagnostic efficiency) and clinical outcomes (survival), providing a replicable model for AMeI care enhancement.

## Discussion

### Quality Improvement Impact on AMeI Outcomes

This study demonstrates that systematic quality improvement in acute mesenteric ischemia care can significantly reduce mortality through measurable process enhancements. Our 2018 quality initiative reduced median emergency department-to-CT time from 8.5 to 2.9 hours, correlating with improved 30-day survival in the endovascular-first cohort from 53.8% to 76.9%. This 23.1 percentage point improvement underscores the critical relationship between diagnostic efficiency and clinical outcomes in time-sensitive vascular emergencies.

The established benchmark of <3 hours for ER-to-CT completion addresses a significant quality gap in AMeI care delivery. While existing literature emphasizes symptom-onset-to-treatment duration, our focus on inhospital time intervals provides actionable targets for healthcare systems. ^8^ The finding that every 6-hour delay doubles mortality risk supports the clinical relevance of these process improvements, yet data quantifying specific diagnostic intervals remain limited across healthcare settings. ^7,11^

### Multidisciplinary Protocol Implementation and Replicability

Our standardized protocol demonstrates the feasibility of implementing evidence-based AMeI care through systematic quality improvement. Key elements include direct specialist activation, mandatory radiological evaluation, and immediate angiography for equivocal cases. The protocol’s evolution from balloon angioplasty to large-bore thrombus aspiration represents technical refinement based on outcome feedback, achieving immediate flow restoration without additional stenting in recent cases.

The multidisciplinary approach addresses care fragmentation, a common barrier in AMeI management. Enhanced team communication, mandatory consultations, and immediate feedback mechanisms contributed to sustained improvement beyond the initial implementation period. This systematic approach provides a replicable framework for other healthcare systems seeking to optimize AMeI outcomes.

### Healthcare Delivery Implications

The contrast between treatment modalities highlights the importance of early intervention strategy selection. While surgery-first achieved 80% 30-day survival, its decline to 40% at one year suggests potential selection bias toward healthier patients or procedural stress in elderly populations. Conservative management’s consistent failure (0% survival) reinforces the necessity of aggressive intervention, even in high-risk patients.

The cost-effectiveness implications of our technical evolution merit consideration. Large-bore aspiration techniques may reduce procedural complexity and device costs while maintaining efficacy, supporting broader protocol adoption. However, the need for specialized equipment and training represents implementation barriers that require institutional commitment and resource allocation.

### Comparison with Contemporary Studies

Our findings align with and extend recent evidence supporting endovascular-first approaches and systematic quality improvement in AMeI care. In a large NSQIP analysis of 439 patients, Branco et al. demonstrated that endovascular therapy was associated with a 2.5-fold decrease in mortality risk compared to open surgery (OR 0.4, P=0.018). ^12^ Our post-2018 endovascular-first survival rate of 76.9% exceeds these national benchmarks, suggesting additional benefits from protocol-driven care delivery beyond treatment modality selection alone.

Contemporary mortality rates from large series remain consistently high across healthcare systems. Open revascularization studies report 30-day mortality rates of 32-38%, while systematic reviews demonstrate overall AMeI mortality of 50-70% despite advances in imaging and intervention techniques. ^13^ Single-center surgical series continue to report mortality rates of 60-80%, particularly when diagnosis is delayed. ^14,15^ In contrast, our quality improvement initiative achieved 23.1% mortality in the endovascular-first cohort post-2018, representing substantial improvement over these contemporary benchmarks.

The concept of “intestinal stroke centers” emphasizing rapid multidisciplinary intervention has gained international recognition, with implementations reported in France and China. ^1,6^ The World Society of Emergency Surgery guidelines emphasize that “introducing clinical pathways and centers of excellence results in higher awareness of AMeI, more appropriate imaging, less delays, an increased number of revascularizations, and therefore lower mortality. ^1^ However, existing literature lacks specific process metrics and measurable quality indicators for implementation. Our <3-hour ER-to-CT benchmark addresses this evidence gap, providing actionable targets that correlate with measurable outcome improvements.

Recent endovascular technique evolution has demonstrated promising results, with studies showing reduced bowel resection rates and shorter hospital stays compared to open approaches. ^16,17^ However, technical success rates vary widely, with reported ranges of 70-90% across different series. ^18,19^ Our technical success rate of 77.8% with zero procedural complications aligns with these reports, while our post-2018 adoption of large-bore thrombus aspiration represents a novel approach not widely documented in current literature. ^20^

Diagnostic delay remains a critical factor across all studies, with Acosta et al. demonstrating that every 6-hour delay doubles mortality risk. ^11^ While previous research has focused on symptom-onset-to-treatment duration, our study is among the first to quantify and optimize specific in-hospital intervals, particularly the emergency department-to-CT pathway. ^7,8^ This focus on measurable process improvement distinguishes our approach from traditional outcome studies and provides a replicable framework for quality enhancement across diverse healthcare settings. ^9^

### Limitations and Generalizability

This single-center registry study reflects real-world practice constraints but may limit generalizability across diverse healthcare settings. The observational design precludes definitive causal inferences, though the temporal relationship between quality improvements and outcome enhancement supports intervention effectiveness. Sample size limitations, particularly in subgroup analyses, require cautious interpretation of findings.

Selection bias and evolving diagnostic criteria over the study period may influence results. The focus on 30-day outcomes, while clinically relevant, may not capture longer-term quality of life or functional outcomes. Additionally, resource requirements for protocol implementation may vary significantly across healthcare systems with different staffing models and technological capabilities.

### Future Research and Implementation

Multicenter validation of the < 3-hour ER-to-CT benchmark is essential for broader adoption. Future research should examine protocol feasibility in resource-limited settings, cost-effectiveness analyses, and optimal implementation strategies across diverse healthcare delivery models. Comparative effectiveness research evaluating different endovascular approaches and hybrid strategies would further refine treatment algorithms.

Quality improvement science methodology could enhance future AMeI protocol development through statistical process control, Plan-Do-Study-Act cycles, and sustainability frameworks. Integration with electronic health records and clinical decision support systems may further optimize care delivery efficiency.

## Conclusion

Systematic quality improvement in acute mesenteric ischemia care demonstrates measurable impact on both process metrics and clinical outcomes. Our decade-long experience shows that reducing diagnostic delays through multidisciplinary protocol implementation significantly improves survival rates. The <3-hour emergency department-to-CT benchmark provides an actionable target for healthcare systems seeking to enhance AMeI care delivery. These findings support the broader application of quality improvement methodology to time-sensitive vascular emergencies, with potential for substantial impact on patient outcomes through systematic care optimization.

## Data Availability

The datasets used and/or analyzed during the current study are available from the corresponding author on reasonable request and subject to institutional review board approval.

## Acknowledgments

Special thanks to the colleagues in the Cardiac Catheterization Laboratory at Hsinchu Branch their assistance in patient care. And we would like to thank Miss Mei-Ching Liao, for her assistance in funding allocation (113-T1-08).

## Author contributions statement

MYH designed the study and performed the analysis. YLL and MYH performed the endovascular treatments.

